# SARS□CoV□2 Associated Shifts in the Upper Respiratory Tract Mycobiome in Non-Hospitalized Cases

**DOI:** 10.64898/2026.05.07.26352639

**Authors:** Siddharth Singh Tomar, Krishna Khairnar

**Author notes:** Corresponding author: Correspondence to Krishna Khairnar.

## Abstract

SARS□CoV□2 infection is associated with marked changes of the upper respiratory tract mycobiome. URT mycobiome Changes in non-hospitalized patients however, remains poorly defined. We performed shotgun metagenomic sequencing of 95 upper respiratory tract swab samples from 48 symptomatic SARS□CoV□2-positive individuals and 47 healthy controls from central India. Fungal diversity and community structure were compared using alpha- and beta-diversity analyses, while differential taxa were identified using prevalence-based testing and LEfSe. SARS□CoV□2-positive samples showed significantly higher fungal alpha diversity than controls, with increased Shannon diversity (p = 0.000319) and Simpson diversity (p = 0.017). Beta-diversity analysis showed significant separation between groups for both Bray–Curtis and Jaccard distances (PERMANOVA p = 0.001), with significant dispersion effects as well (PERMDISP p = 0.001). Differential analysis identified more SARS□CoV□2-enriched than control-enriched taxa, including *Candida orthopsilosis, Malassezia furfur, M. sympodialis, M. globosa, Aspergillus niger, A. terreus,* and *A. nidulans. Aspergillus sydowii* was the main control-enriched taxon. LEfSe and concordant multi-test analysis supported these findings, and sensitivity analysis confirmed robustness across thresholds. Certain SARS□CoV□2-enriched taxa were linked to confirmed or probable COVID□19-associated fungal infections, whereas no such pathogens were detected in controls. These findings indicate that SARS□CoV□2 infection is associated with URT mycobiome dysbiosis and enrichment of clinically relevant opportunistic fungi in community cases.

## Introduction

The emergence of severe acute respiratory syndrome coronavirus 2 (SARS-CoV-2) has posed an unprecedented global health challenge. Clinical outcomes of infected patients range from asymptomatic infection to severe respiratory failure and mortality [1]. While factors such as age, comorbidities, and immune status contribute to disease severity, a growing body of evidence suggests that the respiratory microbiome plays an important role in modulating the outcomes of respiratory viral infections [2,3,4]. The upper respiratory tract (URT), comprising the nasal cavity, nasopharynx, and oropharynx, serves as the primary entry point for SARS-CoV-2 and represents a key site where host–microbe–virus interactions are initiated [5,6].

The URT microbiome regulates the mucosal integrity, prevents pathogen colonization, and maintains ecological balance [7]. Disruption of this equilibrium (dysbiosis) has been reported to increase the susceptibility to respiratory infections and the development of secondary infections [8]. In the context of COVID-19, several studies have reported alterations in the bacterial component of the URT microbiome, including reduced diversity, depletion of commensals, and enrichment of opportunistic pathogens [9,10,11]. However, most of these studies have focused primarily on profiling bacterial communities using 16S rRNA sequencing, thereby providing limited resolution and overlooking other important microbial kingdoms, such as Fungi.

Fungi, which constitute the respiratory mycobiome (the fungal component of the microbiome), represent a critical yet underexplored component of the URT microbiome. The airway mycobiome has been increasingly investigated for its role in shaping host immune responses and influencing disease pathogenesis, particularly in infectious respiratory infections [12,13]. The diversity, ecological dynamics, and functional implications of fungal communities in the URT remained understudied, especially in the context of SARS-CoV-2 infection [14]. Emerging evidence suggests that viral infections can alter the respiratory microenvironment, promoting fungal colonization or overgrowth, thereby increasing the risk of complications, such as secondary fungal infections.

Notably, genera such as *Mucorales*, *Candida*, and *Aspergillus,* which are common constituents of the airway mycobiome, may undergo significant shifts in response to SARS-CoV-2 infection [15,16,17]. These alterations are further aggravated by therapeutic interventions, particularly the widespread use of systemic corticosteroids such as Dexamethasone [18]. Corticosteroids are prescribed to reduce cytokine-mediated hyperinflammation and improve survival in severe COVID-19 cases. Corticosteroids have immunosuppressive effects, thereby creating a permissive environment for fungal colonization and proliferation [19].

Immune suppression combined with comorbidities such as diabetes mellitus and hypertension has been strongly associated with an increased risk of secondary fungal infections. Invasive Fungal Infections associated with COVID-19 are broadly classified as COVID-19-associated pulmonary aspergillosis (CAPA), COVID-19-associated candidiasis (CAC), and COVID-19-associated mucormycosis (CAM) [20]. CAM emerged as a major public health concern during the second wave of COVID-19 in India [21]. CAM carries a high fatality rate if not promptly treated [22]. A systematic review of 20 studies reported a pooled CAPA mortality 51.2%; another meta-analysis of 28 observational studies estimated ICU mortality 54.9% (95% CI 45.6–64.2) [23,24]. CAC is less common but also associated with high mortality; CAC is often linked to central venous lines during ICU admission [25]. Overall, among hospitalized patients, Fungal Coinfection was widely observed to increase the length of hospital stay and the need for mechanical ventilation/ICU care [26].

Shotgun metagenomic sequencing offers a comprehensive, unbiased way to characterize a sample’s microbial community. It captures bacteria, viruses, and fungi in a single assay. Unlike amplicon-based methods, it enables simultaneous profiling of the mycobiome with other microbial components. This supports a systems-level understanding of microbial communities and their links to disease [27]. Huang and colleagues demonstrate the utility of metagenomics for detecting fungal and mixed infections in critically ill COVID-19 patients. Compared with culture- and routine-test-based approaches, metagenomics reveals underrecognized co- and superinfections. It also identifies greater pathogen diversity in SARS-CoV-2 patients [28]. The metagenomic approach is particularly useful for surveillance of differentially abundant pathogenic taxa and their potential links to SARS-CoV-2 infection [29]. It addresses key gaps, including underdiagnosis and incomplete pathogen lists. It improves understanding of host–microbiome interactions and enables faster outbreak detection. However, challenges remain, including the need for standardized interpretation, multicenter validation, and cost-effective implementation.

We have used shotgun metagenomic sequencing to analyze the upper respiratory tract (URT) fungal microbiome of non-hospitalized, symptomatic SARS-CoV-2-positive individuals compared with healthy controls. We have selected community-based cases to elucidate intrinsic modifications in the mycobiome linked to SARS-CoV-2 infection, while reducing confounding influences from hospital-acquired microbial exposure, corticosteroid, and antifungal drug treatment. We specifically aimed to (i) identify differentially abundant fungal taxa associated with SARS-CoV-2 infection, (ii) characterize alterations in fungal community structure, and (iii) assess the potential implications of these changes for respiratory health and disease.

## 2. Materials and Methods

### 2.1 Study Population

Upper respiratory tract (URT) swab samples were collected from individuals in the Vidarbha region of central India during March and April 2023. Participants were recruited from five districts: Nagpur, Wardha, Gadchiroli, Chandrapur, and Bhandara. The study involved two distinct groups. The control group consisted of 48 SARS-CoV-2-negative asymptomatic individuals who tested negative for SARS-CoV-2, Influenza A (H1N1pdm09 and other subtypes), Influenza B, and Respiratory Syncytial Virus (RSV) via reverse-transcription PCR (RT-PCR). The second group included 48 non-hospitalized symptomatic individuals who were SARS-CoV-2-positive and negative for Influenza A (H1N1pdm09 and other subtypes), Influenza B, and Respiratory Syncytial Virus (RSV) via reverse-transcription PCR (RT-PCR). All positive individuals were non-hospitalized and kept in home isolation. Positive individuals were identified as having severe acute respiratory infection (SARI) or influenza-like illness (ILI). The median age was 29 years [IQR 22-37] in the control group and 36 years [IQR 19-67] in the SARS-CoV-2-positive group.

### 2.2 Sample Collection and Processing

A total of 96 upper respiratory tract (URT) swab samples were obtained. 48 samples were from SARS-CoV-2-positive individuals, and 48 were from healthy controls. Samples were collected and stored in viral transport medium (VTM). Trained healthcare professionals performed the collections utilizing standardized nasopharyngeal and oropharyngeal swabbing methods.

Samples were initially stored at 4□°C (≤ 5days) before being transferred to −80□°C for long-term storage. Processing for SARS-CoV-2 RT-PCR was conducted under Biosafety Level-II (BSL-2) conditions. RTPCR cycle threshold (Ct) value of ≤25 for SARS-CoV-2 genes (N, E and RdRP) served as the threshold for positive classification.

### 2.3 DNA Extraction, Library Preparation, and Metagenomic Sequencing

Total DNA was extracted from the upper respiratory tract (URT) sample using the QIAamp DNA Microbiome Kit (Qiagen) according to the manufacturer’s protocols. Concentration of the extracted DNA was assessed using a Qubit fluorometer, and DNA purity was determined using a Nanodrop spectrophotometer (A260/280 and A260/230 ratios). One sample from the control group failed the quality check due to low DNA concentration and was removed from further analysis. Shotgun metagenomic libraries were subsequently prepared using the QIAseq FX DNA Library Preparation Kit. High-throughput paired-end sequencing was performed on the Illumina NextSeq 550 platform using 2×150 bp chemistry. Quality Control of sequencing data was initially performed using FastQC and MultiQC. Per-base Phred quality scores were checked across all reads, and Q30 was used as the principal quality parameter.

### 2.4 Metagenomic Data Analysis

Metagenomic data processing was performed using the Chan Zuckerberg Initiative (CZID) web-based platform [30]. Further processing involved removing External RNA Controls Consortium (ERCC) sequences and filtering adapters, short reads, low-quality sequences, and low-complexity regions using fastp. Sequences with scores below 17, lengths shorter than 35 bp, low-complexity bases exceeding 40%, or more than 15 undetermined bases (Ns) were discarded. To isolate microbial signals, host (human) reads were removed by aligning to the human reference genome. The resulting non-human reads were aligned against the NCBI nucleotide (NT) and protein (NR) databases to generate taxon-specific hit counts via accession-linked annotations. Reads were *de novo* assembled into contigs using SPAdes. Original reads were subsequently mapped back to these contigs to preserve read-contig relationships, followed by BLAST analysis against the NCBI NT and NR databases.

### 2.5 Statistical Analysis and Visualization

Fungal taxon-abundance tables for control and SARS-CoV-2-positive groups were imported from CZID, with taxa normalized into a shared index and (Reads Per Million) RPM-scaled data retained for downstream analysis. Fungal taxa specific to human hosts were filtered using the curated fungal host range database published by Bartoszewicz et al. (2022) [31]. Only taxa with the RPM ≥ 0.5 in at least one group were retained. The RPM threshold is derived by further raising it to a more stringent ≥ 0.5, compared with the recommended ≥ 0.1 by Ghelfenstein-Ferreira et al. (2026) [32]. The samples were annotated by metadata (Group: Control vs SARS-CoV-2). The resulting abundance matrix was used for alpha-diversity and differential-abundance analyses.

Alpha diversity (Shannon and Simpson) and observed□taxon richness were computed per sample and compared between groups using the Mann–Whitney U test. Overall, community composition was visualized via PCA (on post-threshold abundance values) and PCoA (using Bray□Curtis distances), and PERMANOVA was used to test whether fungal community composition differed between SARS□CoV□2 and control groups, and PERMDISP was used to check whether observed separation is due to within-group variability. Jaccard distance was also used along with Bray□Curtis distances. This approach was used to determine whether the observed shifts were predominantly driven by changes in abundance among shared taxa (Bray–Curtis) or by gains and losses of specific fungal taxa across groups (Jaccard).

For differential fungal prevalence, Fisher’s exact test was applied to presence□absence data, and Mann–Whitney U tests were used to compare samples with non-zero RPM values; q□values were computed via the Benjamini–Hochberg (False Discovery Rate) FDR correction. Taxa with a prevalence ratio > 2 and FDR□adjusted p < 0.05 were classified as SARS-CoV-2-enriched, whereas those with a ratio < 0.5 were designated as control□enriched. “Concordant taxa” were identified as a subset that appeared consistently across both Fisher-based prevalence assessments and centered-log-ratio (CLR)-based Mann–Whitney tests. Sensitivity analysis was also performed to examine the stability of enriched□taxon counts across gradients of RPM thresholds and prevalence□ratio thresholds.

To identify fungal taxa most strongly associated with either the SARS-CoV-2 group or the control group, we implemented a LEfSe (Linear Discriminant Analysis (LDA) Effect Size) using relative-abundance tables. Taxon names were transformed into a shared index, and all values were converted to a numeric format with zeros assigned for missing entries. Relative abundance (proportion-based) was used for all tests. For each taxon, the Kruskal–Wallis test and two-sided Mann–Whitney U (Wilcoxon rank-sum) test were calculated between control and SARS-CoV-2 samples. The resulting p-values were adjusted for multiple testing using the Benjamini–Hochberg FDR test. A log-ratio measure (log₂ fold-change) was derived from the mean relative abundances. LDA score was calculated as a weighted sum of −log₁₀ - transformed FDR-adjusted p-values from both tests, signed by the direction of change. Taxa with both Kruskal–Wallis and Wilcoxon FDR-adjusted p-values < 0.05 and an absolute LDA-score ≥ 2.0 were defined as LEfSe-significant fungal taxa.

## 3. Results

### 3.1. Overlap of fungal taxa and filtered abundance patterns

Comparison of raw taxonomic data revealed a shared fungal core comprising 65 taxa common to both the control and SARS-CoV-2 groups. 17 Taxa were control-unique, and 57 were SARS-CoV-2-unique taxa. After applying the RPM ≥ 0.5 filter, the overlap became more selective, resulting in 11 shared taxa, 5 control-unique taxa, and 39 SARS-CoV-2-unique taxa. (**Figure 1a&b)** Heatmap visualization of these RPM-filtered taxa across the 95 samples showed that the SARS□CoV□2 group exhibited a denser, more diverse fungal abundance profile than controls. (**Figure 2)**

**Figure 1:**
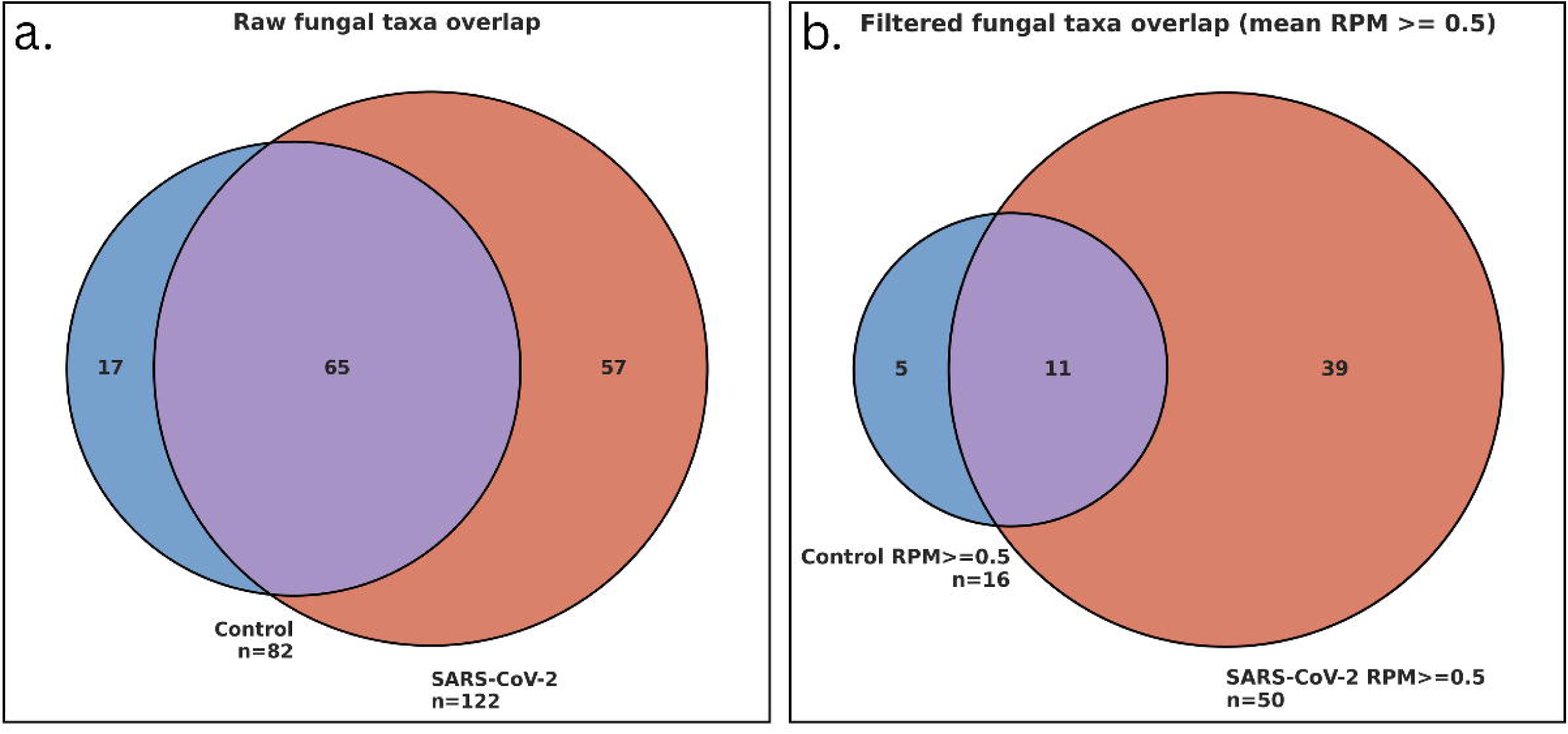
Shared and filtered fungal taxa between SARS-CoV-2 and control groups; (a) Venn diagram showing overlap of raw fungal taxa detected in control and SARS-CoV-2 samples. (b) Venn diagram showing overlap after filtering for RPM ≥ 0.5.

**Figure 2:**
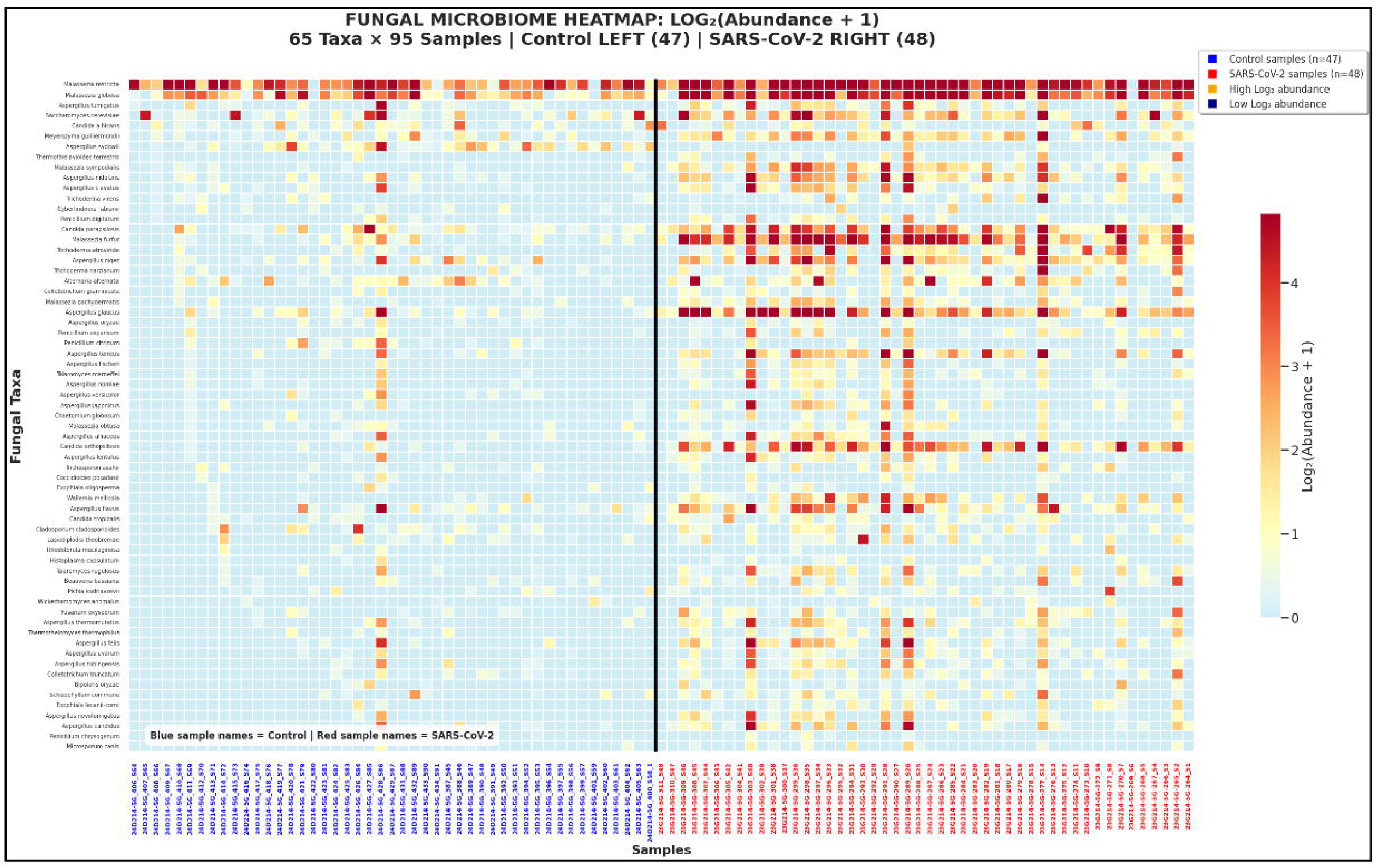
Heatmap of fungal community abundance across control and SARS□CoV□2 samples; Normalized fungal abundance values (log2[abundance + 1]) are displayed for taxa detected across samples, with control samples shown on the left and SARS□CoV□2 samples on the right.

### 3.2. Alpha and Beta Diversity Analysis

Fungal alpha diversity was significantly altered in the SARS□CoV□2 group, with higher Shannon diversity than in controls (Mann–Whitney p = 0.000319). Simpson diversity also differed significantly between groups (Mann–Whitney p = 0.017). (**Figure 3a&b)** Beta-diversity analyses demonstrated significant separation between groups for both Bray–Curtis and Jaccard distances (PERMANOVA p = 0.001 for each), and PERMDISP was also significant for both metrics (p = 0.001). PERMANOVA indicated a significant difference in fungal community composition between groups, while PERMDISP showed that group dispersion also differed, suggesting that both compositional shifts and variability contributed to the observed separation. (**Figure 3c&d)**

**Figure 3:**
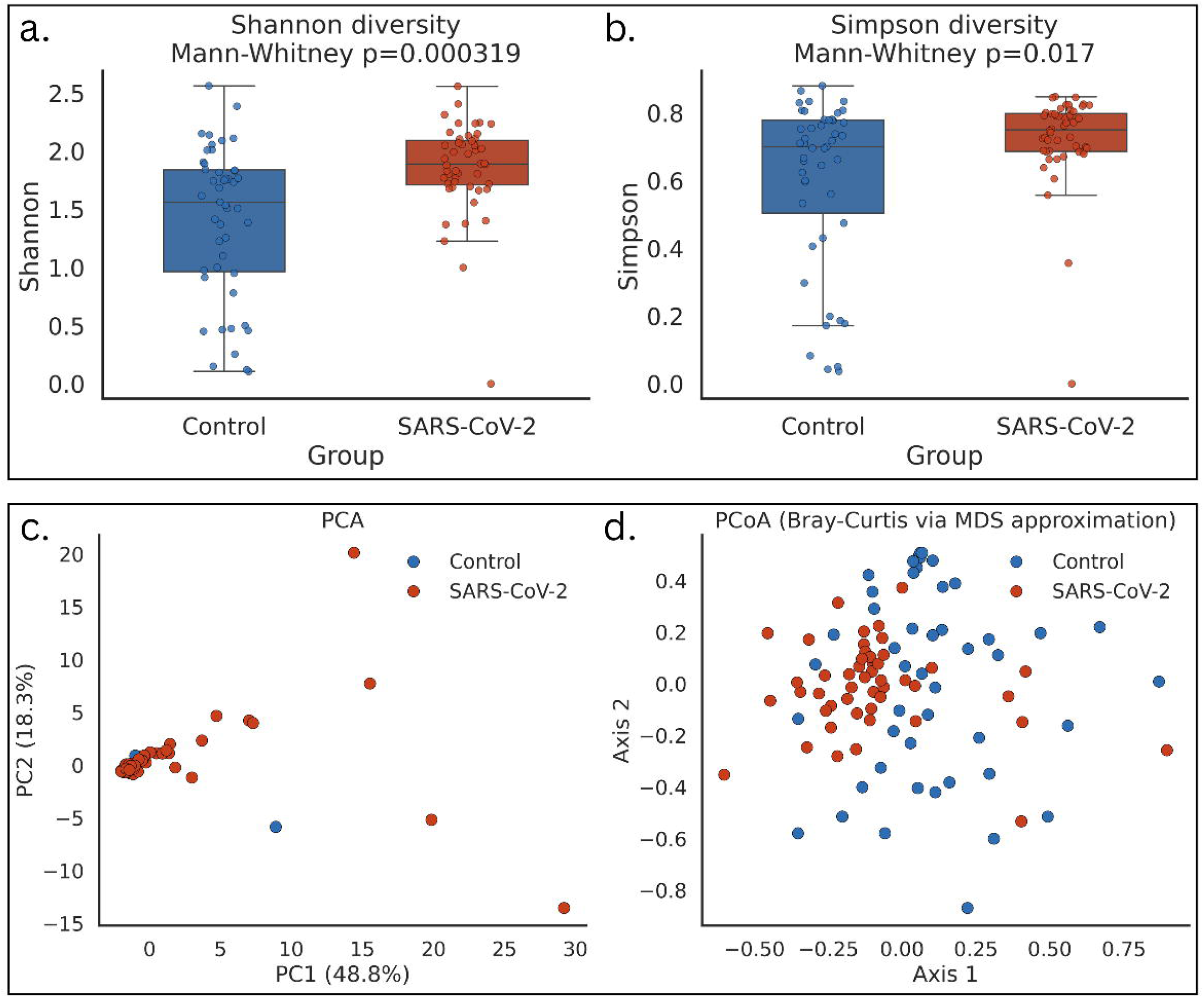
(a) Shannon diversity differed significantly between groups (Mann–Whitney p = 0.000319), with higher diversity in SARS□CoV□2 samples. (b) Simpson diversity also differed significantly (Mann–Whitney p = 0.017). (c) PCA and (d) PCoA based on Bray–Curtis distances showed separation between SARS□CoV□2 and control samples.

### 3.3. Differentially Enriched Fungal Taxa

Differential testing compared fungal taxa between SARS□CoV□2-positive and control samples using Fisher’s exact test for prevalence. Mann–Whitney testing was applied to positive-only abundance, and CLR-based Wilcoxon testing was performed to confirm that observed taxon-level differences were robust to the dataset’s relative-abundance structure. Multiple-testing correction was applied, and taxa with consistent directionality (SARS-CoV-2 or Control) across tests were classified as concordant. This approach showed a higher number of SARS□CoV□2-enriched taxa than control-enriched taxa, with *Candida orthopsilosis*, *Malassezia sympodialis, Malassezia furfur*, and *Aspergillus glaucus* among the top SARS-CoV-2-enriched taxa, while *Aspergillus sydowii* was the only control-enriched taxon. Detailed results are provided in **(Supplementary data/Table S1).** The volcano plot showed that the most significantly differentiated taxa were shifted toward the SARS□CoV□2-associated side, exceeding both the FDR-adjusted P-value and prevalence-ratio thresholds. (**Figure 4)**

**Figure 4:**
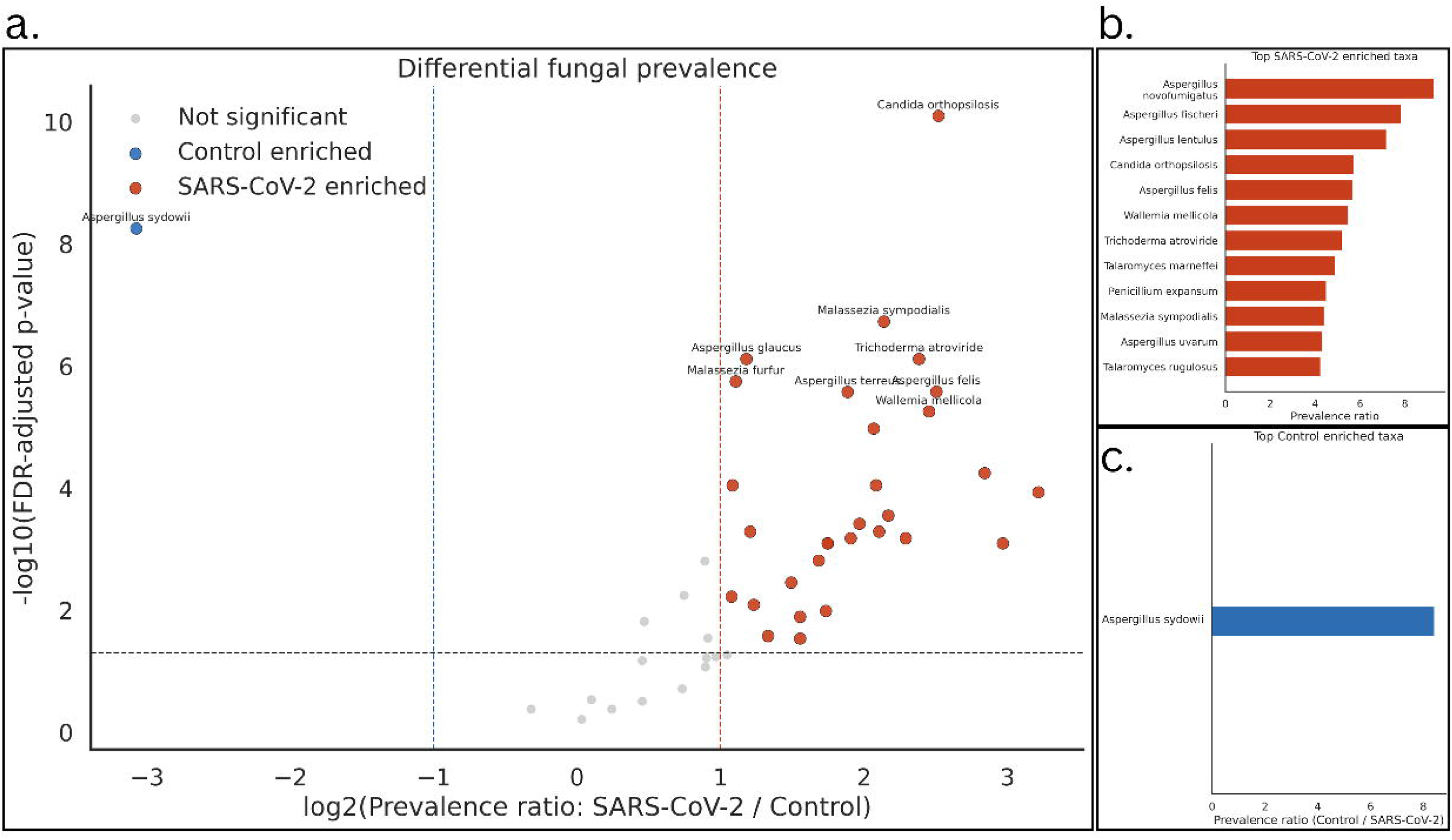
(a) Volcano plot showing differential prevalence based on log2 prevalence ratio and FDR-adjusted significance. Taxa enriched in SARS□CoV□2 are shown in red, control-enriched taxa in blue, and non-significant taxa in gray. (b) Top SARS□CoV□2–enriched taxa ranked by prevalence ratio. (c) Top control-enriched taxa ranked by prevalence ratio.

### 3.4. LEfSe-Based Identification of Differential Fungal Taxa

The most differentially abundant taxa with the highest LDA scores included *Candida orthopsilosis*, *Malassezia furfur*, *Aspergillus glaucus*, and *Malassezia sympodialis*, followed by *Fusarium vanettenii, Aspergillus terreus,* and *Trichoderma atroviride*. Additional SARS-CoV-2-associated taxa comprised *Malassezia globosa, Aspergillus niger, Wallemia mellicola, and Candida parapsilosis*. In contrast, only a limited number of taxa, including *Malassezia arunalokei*, *Aspergillus chevalieri*, and *Aspergillus sydowii*, were enriched in control samples, with comparatively lower representation. Overall, the LDA score distribution showed a stronger, more diverse discriminatory signal in SARS-CoV-2 samples. (**Figure 5)**

**Figure 5:**
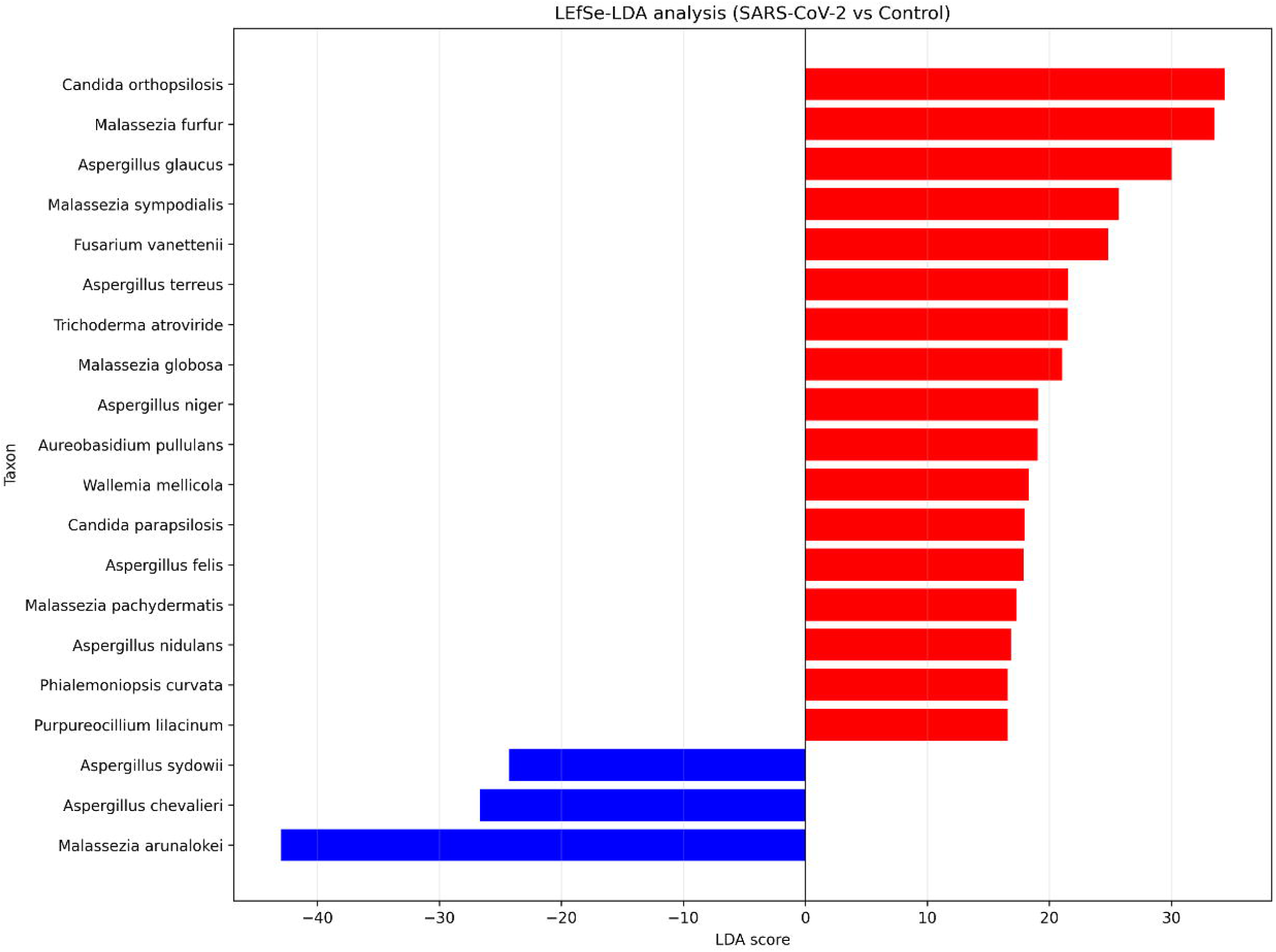
Linear discriminant analysis (LDA) effect size (LEfSe) identified taxa enriched in the SARS□CoV□2 group (red) and control group (blue). Bars represent LDA scores; higher scores indicate stronger group association.

### 3.5. Robustness Across Threshold Settings

Sensitivity analysis showed that the overall differential enrichment pattern persisted across gradients of RPM and prevalence-ratio thresholds. While the number of significant SARS□CoV□2-enriched taxa decreased as thresholds became more stringent, the enrichment pattern persisted throughout the tested threshold gradient. Control-enriched taxa remained less abundant across all threshold ranges. This indicates that the principal findings were not dependent on a single cutoff choice. (**Figure 6)**

**Figure 6:**
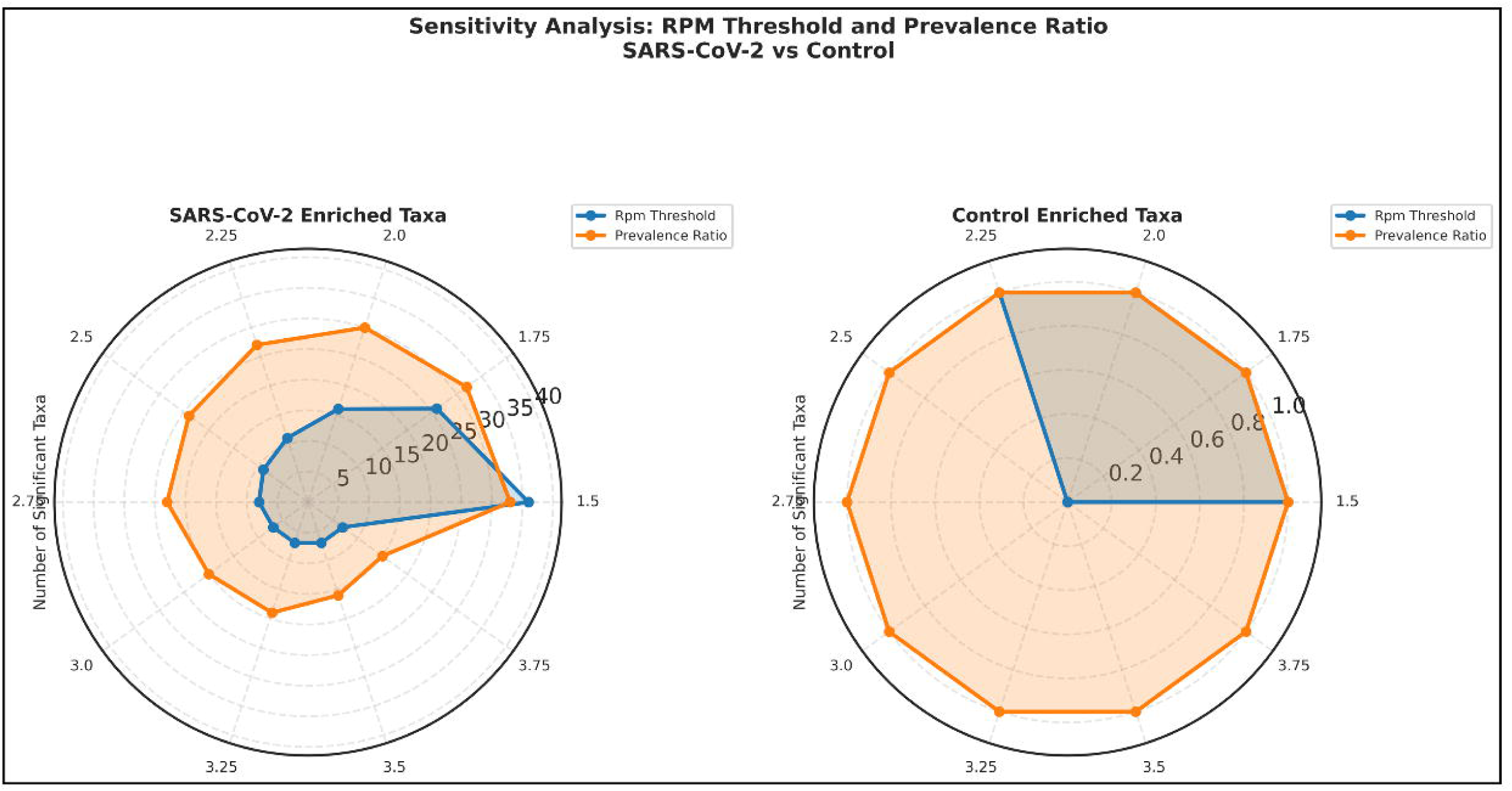
Radar plots compare the number of significant taxa identified across different RPM thresholds and prevalence-ratio cutoffs for SARS□CoV□2-enriched and control-enriched taxa.

### 3.6. Pathogenic Burden in SARS-CoV-2 and Control Group

The SARS□CoV□2 group was observed to be enriched with fungal taxa linked to clinical secondary infections. LEfSe significant taxa were classified into **Confirmed**, **Probable**, **Limited**, and **Not reported** categories. This classification was based on the availability of clinical studies of COVID□19 associated secondary fungal infections pertaining to a particular taxon. Confirmed taxa included *Aspergillus niger, A. terreus, A. nidulans, Candida parapsilosis*, and *Meyerozyma guilliermondii*. These are reported in CAPA, candidemia, or invasive candidiasis in hospitalized patients. Probable taxa included *Aspergillus felis, Candida orthopsilosis*, and *Malassezia* species *M. furfur, M. globosa*, and *M. sympodialis*. These are consistent with opportunistic infection or folliculitis in COVID□19 settings. The control group had no taxa classified as confirmed or probable pathogens. (**Table 1)**

**Table 1:** Fungal taxa identified by LEfSe in SARS□CoV□2 and control groups, with fungal classification, enrichment direction, evidence level for COVID□19 association, reported infection type, and corresponding references.

| Taxon | Fungal Classification | LEfSe Enriched Group | Evidence level | Reported Infection | References |
| --- | --- | --- | --- | --- | --- |
| *Aspergillus niger* | Mould | SARS-CoV-2 | Confirmed | Pulmonary aspergillosis (CAPA), rhinosinusitis | 43, 48 |
| *Aspergillus terreus* | Mould | SARS-CoV-2 | Confirmed | Pulmonary aspergillosis (CAPA) | 43, 49 |
| *Aspergillus nidulans* | Mould | SARS-CoV-2 | Confirmed | Pulmonary aspergillosis (CAPA) | 43 |
| *Aspergillus felis* | Mould (cryptic, section Fumigati) | SARS-CoV-2 | Probable | Pulmonary aspergillosis | 20,49 |
| *Aspergillus glaucus* | Mould | SARS-CoV-2 | Limited | Opportunistic infection | No COVID-19-specific reports found at the species level |
| *Aspergillus chevalieri* | Mould | Control | Not reported | — | No COVID-19-specific reports found at the species level |
| *Aspergillus sydowii* | Mould | Control | Limited | Opportunistic infection | 46 |
| *Candida parapsilosis* | Yeast | SARS-CoV-2 | Confirmed | Candidemia, invasive candidiasis | 38, 49 |
| *Candida orthopsilosis* (Part of *C. parapsilosis* complex) | Yeast | SARS-CoV-2 | Probable | Candidemia | 38, 49 |
| *Meyerozyma guilliermondii* | Yeast | SARS-CoV-2 | Confirmed | Invasive candidiasis, candidemia | 50 |
| *Malassezia furfur* | Yeast (basidiomycete) | SARS-CoV-2 | Probable | Malassezia folliculitis | 41 |
| *Malassezia globosa* | Yeast (basidiomycete) | SARS-CoV-2 | Probable | Malassezia folliculitis | 41 |
| *Malassezia sympodialis* | Yeast (basidiomycete) | SARS-CoV-2 | Probable | Malassezia folliculitis | No COVID-19-specific reports found at the species level |
| *Malassezia pachydermatis* | Yeast (basidiomycete) | SARS-CoV-2 | Limited | Opportunistic fungemia | No COVID-19-specific reports found at the species level |
| *Malassezia arunalokei* | Yeast (basidiomycete) | Control | Not reported | — | No COVID-19-specific reports found at the species level |
| *Fusarium vanettenii* | Mould | SARS-CoV-2 | Limited | Invasive fusariosis | No COVID-19-specific reports found at the species level |
| *Trichoderma atroviride* | Mould | SARS-CoV-2 | Limited | Invasive fungal rhinosinusitis | No COVID-19-specific reports found at the species level |
| *Purpureocillium lilacinum* | Mould | SARS-CoV-2 | Limited | Opportunistic hyalohyphomycosis | No COVID-19-specific reports found at the species level |
| *Phialemoniopsis curvata* | Mould | SARS-CoV-2 | Not reported | — | No COVID-19-specific reports found at the species level |
| *Wallemia mellicola* | Basidiomycete (xerophilic) | SARS-CoV-2 | Not reported | — | No COVID-19-specific reports found at the species level |

## 4. Discussion

This study provides one of the first shotgun metagenomic profiles of the upper respiratory tract (URT) mycobiome in non-hospitalized, community-based SARS-CoV-2-positive patients. Hospitalized patients were excluded to remove the confounding effects of corticosteroid therapy, antifungal prophylaxis, broad-spectrum antibiotics, and nosocomial exposure. Hospitalization-induced changes are widely reported in the COVID-19 mycobiome literature [20,33]. The resulting mycobiome could thereby be more representative of the intrinsic changes of the URT fungal community. offering insight into ecological changes that may precede or predispose to more severe downstream fungal complications.

Significantly elevated fungal alpha diversity and clear beta-diversity separation between SARS-CoV-2-positive and control groups indicate remodeling of the URT mycobiome. These observations align and further the prior metatranscriptomic work by Hoque et al. (2022); the authors have demonstrated significantly higher fungal diversity and greater abundance of opportunistic pathogens in the nasopharyngeal tract of COVID-19 patients compared with healthy and recovered controls [34].

The observation of significantly higher Shannon (p = 0.000319) and Simpson (p = 0.017) alpha diversity indices in SARS-CoV-2-positive individuals relative to controls is a major finding of our study. This finding should be interpreted in the specific context of respiratory microbiome research. In the respiratory microbiome, elevated diversity does not necessarily indicate a healthy or resilient microbial community; rather, it can signal dysbiotic expansion of normally low-abundance or intermediate taxa that may exploit a disrupted mucosal environment. The pattern of increased URT microbiome diversity has been reported in multiple studies [34,35,36].

The divergence in beta-diversity, confirmed by PERMANOVA significance (p = 0.001) for both Bray–Curtis and Jaccard distances alongside significant PERMDISP results, indicates that the mycobiome shifts reflect both compositional alterations (gains and losses of specific taxa) and differential abundance of shared taxa. This finding suggests that SARS-CoV-2 infection not only makes it conducive for new taxa to establish themselves but simultaneously promotes the overgrowth of taxa already present in low threshold abundances. This is consistent with the ecological ‘Anna Karenina principle,’ in which host stress destabilizes microbiomes and increases random variation [37].

The concordant taxa identified across Fisher’s exact test, Mann–Whitney U test, CLR-based Wilcoxon test, and LEfSe analysis provide a multi-test-supported list of taxa. Sensitivity analysis across a gradient of abundance and prevalence ratio thresholds was used to evaluate statistical stability. This ruled out the possibility that the observed alterations in URT prevalence or abundance were merely artifacts of threshold selection. (**Figure 6)**.

*Candida orthopsilosis* is the top-ranked SARS-CoV-2-enriched taxon by LDA score. *C. orthopsilosis* is a member of the *C. parapsilosis* species complex comprising *C. parapsilosis, C. orthopsilosis,* and *C. metapsilosis* [38]. The COVID-19 pandemic was associated with a marked increase in *C. parapsilosis* candidemia, with outbreaks of fluconazole-resistant fungus reported across European hospitals [39]. Enrichment of *C. orthopsilosis* in the URT of non-hospitalized COVID-19 patients in the current study suggests that mucosal colonization by this complex could be established early in infection and may serve as a starting point for subsequent invasive infection.

*Malassezia furfur, M. sympodialis,* and *M. globosa* were also enriched in SARS-CoV-2-positive individuals. *Malassezia* is one of the most abundant fungi in the human skin and mucosal microbiome and is generally classified as normal skin flora [40]. In the context of COVID-19, Malassezia folliculitis has been described in hospitalized patients receiving systemic dexamethasone, with the corticosteroid implicated in increasing sebum production, thereby increasing the abundance of lipid-dependent *Malassezia* [41]. However, the enrichment observed here occurred in non-hospitalized patients without corticosteroid exposure, warranting further investigation into its mechanistic basis. We propose a plausible mechanism for this finding, given that SARS-CoV-2 infection impairs the Type 17 helper cell (Th17) immune axis [33], which is a principal determinant of mucosal antifungal defense. Th17 produces interleukin (IL-17), which mediates immune responses that control *Malassezia* colonization of the skin and mucosal tissues. An impaired Th17 axis could lead to pathological *Malassezia* overgrowth [42].

The enrichment of *Aspergillus niger, A. terreus, A. nidulans,* and *A. glaucus* in SARS-CoV-2-positive participants is converging with a growing body of evidence that *Aspergillus* colonization precedes invasive pulmonary aspergillosis (IPA). In a prospective surveillance study of 122 COVID-19 patients, Ogawa et al. (2023) cultured *A. nidulans, A. niger,* and *A. terreus* from respiratory samples alongside *A. fumigatus* [43].

The detection of *Aspergillus spp.* in the URT of community-based, non-hospitalized patients suggests that airway colonization by pathogenic *Aspergillus spp*. may be established before hospitalization and independently of intensive care interventions. This observation supports the hypothesis that COVID-19 itself, through direct epithelial damage and immunological impairment, promotes conditions favorable to Aspergillus colonization.

The enrichment of *Fusarium vanettenii* and *Trichoderma atroviride* in SARS-CoV-2-positive individuals is consistent with broader reports of emerging hyalohyphomycosis infections in COVID-19 patients [44]. At the genus level, *Fusarium spp*. have been reported as causative agents of invasive fungal rhinosinusitis in COVID-19 patients [45].

The finding that the control group harbored no ‘Confirmed’ or ‘Probable’ pathogenic taxa, while the SARS-CoV-2-positive group was enriched with multiple such organisms. It implies that URT metagenomics could, in principle, serve as a surveillance tool for identifying individuals at elevated risk of secondary infection during early COVID-19 infection. The control-enriched taxa *Aspergillus chevalieri, A. sydowii,* and *Malassezia arunalokei* are xerophilic fungi, generally classified as taxa of limited pathogenic significance. While *A. sydowii* has recently been documented as an occasional opportunistic pathogen in immunocompromised patients in a case report [46], its consistent enrichment in controls rather than COVID-19 patients in this cohort suggests a commensal prevalence role in the healthy URT microenvironment.

Our Study suggests that mycobiome-based risk classification could be integrated into early COVID-19 management, for example, patients with detectable enrichment of *C. parapsilosis, Aspergillus spp.,* or *Malassezia spp*. at initial presentation could be flagged for heightened antifungal surveillance should their disease progress. This is particularly relevant in high-burden settings such as India. The findings also underscore the value of metagenomic surveillance as a complementary tool to culture-based diagnostics. Compared with conventional methods, metagenomics identifies a broader spectrum of fungal pathogens, including rare and low-abundance taxa [46]. The integration of rapid metagenomics-based testing into clinical algorithms for COVID-19 patients, particularly those presenting with ILI/SARI, could reduce diagnostic delays. The sensitivity analysis demonstrating consistent enrichment patterns across a range of RPM and prevalence-ratio thresholds enhances confidence in the robustness of the differential taxon list and supports its potential application to future large-scale surveillance datasets.

## Conclusions and Future Prospects

We have demonstrated that SARS-CoV-2 infection in non-hospitalized, community-based individuals could lead to significant alterations of the URT mycobiome. These alterations are characterized by elevated fungal diversity and enrichment of clinically significant opportunistic taxa. The absence of corticosteroid or antifungal exposure in the groups helps establish the contribution of SARS-CoV-2 infection to the observed mycobiome alterations. Our findings suggest that SARS-CoV-2-driven immune dysregulation and alterations in the URT microenvironment alone are sufficient to create a permissive URT environment for opportunistic fungal colonization.

Future research should focus on longitudinal mycobiome profiling across COVID-19 severity levels. It is also important to integrate culture-based and molecular confirmation of key enriched taxa. Large multicenter studies should account for geographic, demographic, and variant-specific variables. Addressing the gap between mycobiome surveillance and clinically actionable insights will require both methodological optimization and investment in diagnostic infrastructure in resource-limited settings.

## Limitations

The study did not subclassify patients by symptom severity. Severity data were unavailable at the time of collection under the original surveillance mandate. Details on antibiotic/antifungal use were also unavailable, as these were non-hospitalized community cases. Such factors may modulate the magnitude of immune disruption and mycobiome alteration. Additionally, shotgun metagenomics detects fungal DNA, but symptomatic corroboration is required to ascertain the active infections.

## Author declaration

The authors assure that the research has complied with all ethical guidelines and has received approval from the Institutional Ethics Committee for Research on Human Subjects (IEC) of CSIR-NEERI, Nagpur-20, India. Necessary consent from patients/participants has been obtained, and relevant institutional documentation has been archived. The Manuscript is approved by the Institute’s Knowledge Resource Center (KRC) **CSIR-NEERI/KRC/2026/MAY/EEPMU/1**

## Confidentiality declaration

Sample IDs (23G214-5G-264_S1 to 23G214-5G-311_S48 and 24D214-5G_387_S45 to 24D214-5G_434_S91) are masked IDs and cannot be traced to participant details.

## Author contribution statement

SST and KK have contributed equally to the conceptualization, experimentation, and data analysis of this study.

## Conflict of interest statement

The authors declare no conflict of interest

## Data Availability

All data produced in the present study are available upon reasonable request to the authors

## Acknowledgement

The authors are thankful to CSIR-NEERI for providing funds under project OLP-57 (March 2023 -April 2024) for conducting this study

## Notes

### Competing Interest Statement

The authors have declared no competing interest.

### Author Declarations

Institutional Ethics Committee for Research on Human Subjects (IEC) of CSIR-NEERI, Nagpur-20, India

